# Maternal cell-free RNA versus combined screening for first-trimester prediction of early-onset preeclampsia: a nested case-control study

**DOI:** 10.64898/2026.08.28.26361628

**Authors:** Elena Satorres-Pérez, Nerea Castillo-Marco, Marina Igual, Teresa Cordero, Irene Muñoz-Blat, Rogelio Monfort-Ortiz, Beatriz Marcos-Puig, Carlos Simón, Tamara Garrido-Gómez, Alfredo Perales-Marín

## Abstract

**Background:** In Europe, first-trimester combined screening with the Fetal Medicine Foundation (FMF) algorithm identifies women at increased risk of preeclampsia who may benefit from personalized aspirin prophylaxis. However, a substantial proportion of early-onset preeclampsia (EOPE) remains undetected at clinically acceptable specificity.

**Objective:** To evaluate the first-trimester performance of MaiRa for early-onset preeclampsia (EOPE) risk stratification by benchmarking it against FMF screening in the same women, characterizing discordant patient-level classification profiles and exploring potential implementation strategies.

**Study Design:** This secondary case-control analysis was nested within the prospective, multicentre PREMOM cohort [NCT04990141], which enrolled women with singleton pregnancies across 14 tertiary hospitals in Spain. First-trimester MaiRa and FMF risk estimates were evaluated in the same 126 pregnant women, comprising 99 uncomplicated controls and 27 EOPE cases, defined by disease onset before 34 weeks. Discrimination was compared using a stratified paired bootstrap analysis of the areas under the receiver-operating-characteristic curves. Performance was assessed at prespecified clinical thresholds, and detection rates were evaluated at fixed false-positive rates. Universal and contingent MaiRa implementation strategies were also evaluated.

**Results:** MaiRa showed greater first-trimester discrimination for EOPE than FMF combined screening (AUC, 0.974 vs 0.900; *P*=.040) and consistently achieved higher detection rates across fixed false-positive rates. At false-positive rates of 5% and 10%, MaiRa detected 85.2% and 92.6% of EOPE cases, compared with 44.4% and 70.4% for FMF, respectively. Patient-level analysis demonstrated that MaiRa identified 12 of 27 EOPE cases (44.4%) classified as low risk by FMF; these pregnancies generally exhibited less abnormal conventional first-trimester profiles, including fewer maternal risk factors, lower mean arterial pressure and lower uterine artery pulsatility index, yet 8 of 12 (66.7%) subsequently developed severe EOPE. Exploratory implementation analyses showed that universal MaiRa screening achieved the highest EOPE detection, whereas a contingent strategy using FMF for triage and reflex MaiRa testing reduced molecular testing to 35.7% of pregnancies while maintaining 77.8% sensitivity and 97.0% specificity.

**Conclusion:** MaiRa provided greater first-trimester discrimination for EOPE than conventional combined screening and detected additional pregnancies that later developed severe disease despite less abnormal conventional screening profiles. The findings suggest that maternal plasma cfRNA profiling captures biological alterations not fully reflected by combined first-trimester screening and support further prospective evaluation in an independent, unselected obstetric population.

**CONDENSATION:** *Tweetable Statement:* MaiRa outperformed combined screening for early-onset preeclampsia and identified severe cases with less abnormal conventional risk profiles, supporting the added clinical value of molecular screening.

*AJOG at a Glance:*

**A. Why was this study conducted?**

- Combined first-trimester screening identifies women eligible for aspirin prophylaxis but leaves a proportion of early-onset preeclampsia at clinically acceptable specificity.
- In the same individuals, we evaluated whether MaiRa, a test derived from maternal cell-free RNA, could improve first-trimester prediction and assessed strategies for its clinical implementation.
**B. What are the key findings?**

- At a fixed 10% false-positive rate, MaiRa detected 92.6% of early-onset preeclampsia cases compared with 70.4% for combined screening.
- It also identified additional severe cases with less abnormal conventional risk profiles, supporting cfRNA profiling to capture the biological complexity of early-onset preeclampsia.
- Universal MaiRa screening achieved the highest detection, whereas a contingent strategy reduced molecular testing while maintaining high performance.
**C. What does this study add to what is known?**

- Molecular cfRNA-based risk assessment can identify early-onset preeclampsia cases missed by combined screening and informs translation into clinical pathways.

## Introduction

Preeclampsia remains a major cause of maternal and perinatal morbidity and mortality.^1,2^ Early identification of women at risk is essential to enable preventive strategies and closer surveillance, including timely, personalized initiation of aspirin prophylaxis before 16 weeks.^3–6^ Because maternal characteristics and medical history alone provide limited predictive performance, more comprehensive first-trimester screening strategies have been developed.^7,8^ The Fetal Medicine Foundation (FMF) algorithm^7,8^ combines maternal characteristics with biophysical and biochemical markers—most commonly mean arterial pressure, uterine artery Doppler and placental growth factor— and improves detection of preterm preeclampsia (defined as delivery before 37 weeks) relative to history-based screening.^9–12^ However, its performance varies with the markers included, their measurement quality, local implementation, and the risk threshold applied,^11–13^ and higher detection is generally achieved at the expense of specificity. At a fixed 10% false-positive rate, first-trimester combined screening detects approximately 75% of preterm preeclampsia cases and up to 90% of cases resulting in delivery before 34 weeks,^11^ a subgroup generally enriched for more severe disease.^14^ Even so, a clinically relevant proportion of cases remains classified as low risk at standard thresholds.

This limitation is particularly relevant for early-onset preeclampsia (EOPE), defined by disease onset before 34 weeks^15^ and associated with the greatest maternal and perinatal morbidity.^16–20^ EOPE is preceded by early disturbances in maternal adaptation to pregnancy, including defective decidual function,^21–24^ immune dysregulation,^25,26^ and impaired vascular remodelling^27^—that arise before overt placental dysfunction becomes clinically apparent.^28–30^ Because these early molecular alterations may not yet be captured by conventional first-trimester maternal characteristics or biophysical and biochemical markers, transcriptomic profiling offers the opportunity to interrogate biological processes that current screening does not directly assess.

Maternal plasma cell-free RNA (cfRNA) has recently emerged as a means of capturing ongoing maternal and placental processes during pregnancy.^31–34^ By integrating dynamic transcriptomic signals from maternal and placental tissues—spanning decidual function, immune adaptation, vascular remodeling, and placental development— cfRNA provides a direct molecular readout of the biological processes underlying early disease. Building on this principle, MaiRa was developed as a maternal cfRNA-based molecular classifier for first-trimester prediction of EOPE.^35^ Whether this molecular approach improves EOPE risk stratification compared with conventional FMF combined screening has not been directly assessed.

In this study, we evaluated MaiRa as a first-trimester molecular screening test for EOPE, using the FMF combined-screening model as the clinical comparator in the same individuals. We compared discriminatory performance, examined patient-level classification profiles to determine whether both approaches identified the same or different EOPE pregnancies, and explored potential clinical implementation strategies for molecular screening.

## Materials and Methods

### Study design and population

This was a secondary case-control analysis nested within the prospective, longitudinal PREMOM cohort, which enrolled women with singleton pregnancies between 9 and 14 weeks of gestation across 14 tertiary hospitals in Spain (ClinicalTrials.gov identifier NCT04990141).^35^ This multicenter design captures population diversity and between-centre variability in routine first-trimester screening. For the present comparative analysis, pregnancies were eligible if first-trimester blood sampling and FMF assessment were performed within the 11+0 to 13+6-week window and both a FMF risk estimate and a MaiRa result were available. The final analysis included 126 pregnancies, comprising 99 uncomplicated controls and 27 EOPE cases. EOPE was defined as preeclampsia with disease onset before 34 weeks.^15^

### FMF and MaiRa preeclampsia risk assessment

For each pregnancy, the PREMOM database recorded a first-trimester FMF risk estimate and a first-trimester MaiRa score. FMF risk was calculated using the official preeclampsia algorithm, integrating maternal characteristics, mean arterial pressure, uterine artery pulsatility index, placental growth factor, and pregnancy-associated plasma protein-A; the patient-specific risk was expressed as 1/N (lower denominators indicating higher risk) and converted to the corresponding probability for continuous analyses. MaiRa provided a continuous score generated from maternal plasma cfRNA using the previously developed first-trimester EOPE model,^35^ with higher values indicating greater predicted risk. For thresholds-based analyses, high risk was prespecified as ≤1/100 for FMF and ≥0.251 for MaiRa. For comparisons at fixed false-positive rates, detection rates were estimated for FMF and MaiRa at false-positive rates of 5%, 10%, 15%, and 20%.

### Clinical-use scenarios

Three clinical-use strategies were evaluated: FMF standalone, MaiRa standalone, and contingent MaiRa testing. FMF standalone used the prespecified high-risk threshold of ≤1/100, and MaiRa standalone classified all pregnancies at ≥0.251. In the contingent strategy, FMF served as the initial triage test and the MaiRa threshold was fixed at ≥0.251 (pregnancies meeting the FMF triage criterion underwent reflex MaiRa testing and were classified as high risk when the MaiRa score was ≥0.251, whereas those not meeting the criterion were classified as low risk without molecular testing). The FMF triage cutoff was progressively broadened, and sensitivity, specificity, and the proportion of pregnancies requiring MaiRa testing were calculated for each cutoff; a cutoff of ≤1/300 was selected for the primary contingent analysis (Supplementary Figure 1).

### Statistical analysis

Continuous variables are presented as medians with interquartile ranges, and categorical variables as counts with percentages. Groups were compared using Welch’s t test for approximately normally distributed continuous variables, the Mann–Whitney U test for non-normally distributed continuous variables, and the Fisher exact test for categorical variables. Discrimination for EOPE was assessed using receiver-operating-characteristic curves and quantified by the area under the curve (AUC). MaiRa and FMF AUCs were compared within the same pregnancies using a stratified paired bootstrap procedure with 10,000 resamples. Detection rates and 95% confidence intervals were estimated for each method at fixed false-positive rates of 5%, 10%, 15%, and 20%. For threshold- and strategy-based analyses, sensitivity, specificity, and the number and proportion of pregnancies requiring MaiRa testing were calculated. Agreement between the binary MaiRa and FMF risk classifications was assessed using Cohen’s κ coefficient with its 95% confidence interval, and the strength of agreement was interpreted according to the Landis and Koch criteria.^36^ All tests were 2-sided, and P<.05 was considered statistically significant.

## Results

### Study population

The nested case-control analysis included 126 singleton pregnancies from the PREMOM cohort^35^—99 uncomplicated controls and 27 that subsequently developed EOPE—each with maternal plasma collected at 11 to 13 weeks and complete data for both MaiRa and conventional first-trimester combined screening. This cohort provided the basis for a head-to-head comparison of MaiRa and conventional first-trimester combined screening, including analyses of diagnostic performance, patient-level classification, and potential implementation models (Figure 1). Maternal characteristics, first-trimester clinical variables, and pregnancy outcomes are summarized in Table 1.

**Figure 1.**
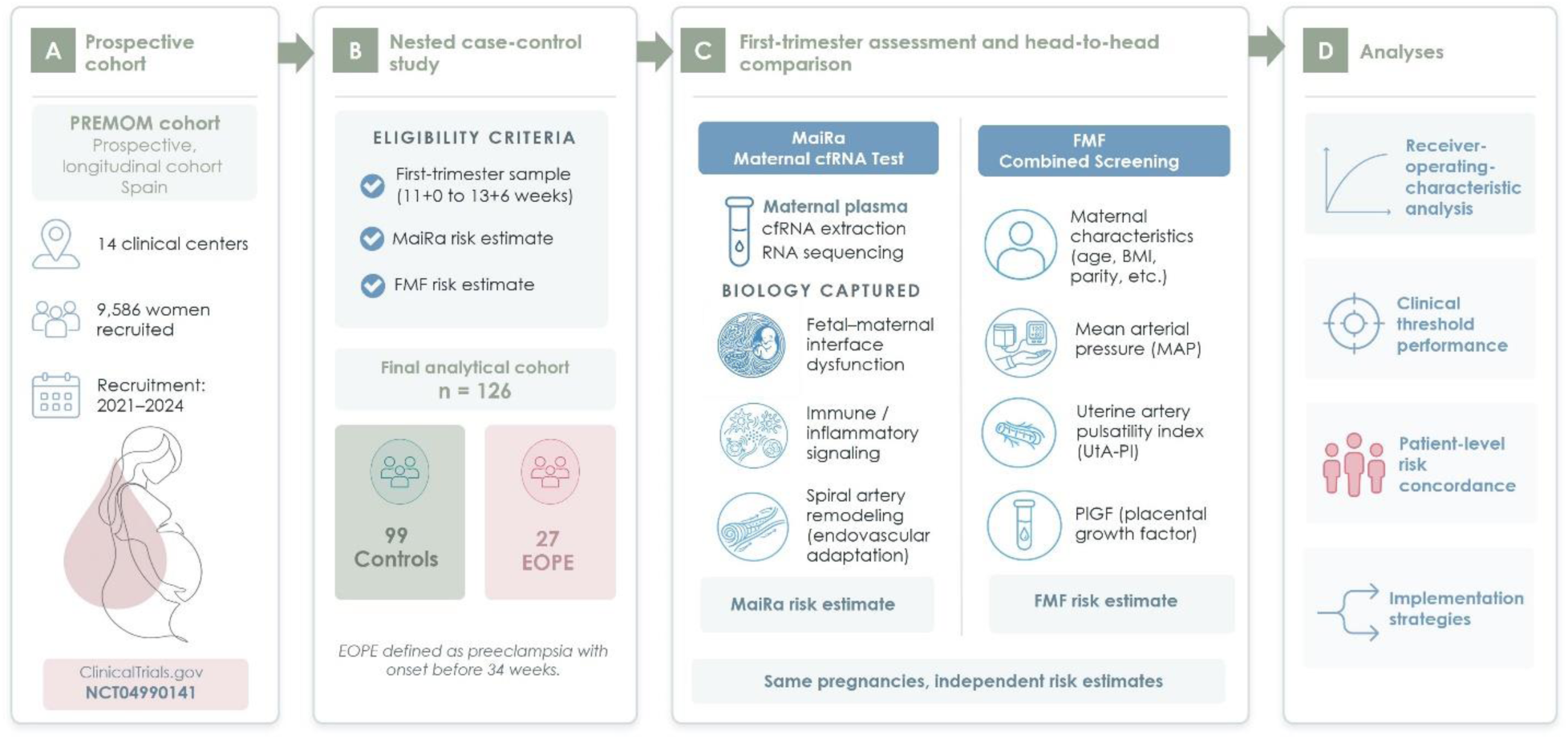
Overview of the study design and the analytical framework used to compare MaiRa and conventional first-trimester combined screening. The analysis included 126 singleton pregnancies, comprising 27 EOPE cases and 99 controls, selected from the prospective PREMOM cohort. (A) Overview of the prospective PREMOM cohort. (B) Eligibility criteria for inclusion in the nested case-control analysis. (C) Composition of the final analytical cohort. (D) Head-to-head first-trimester assessment comparing FMF combined screening and MaiRa performed in the same pregnancies. FMF combines maternal characteristics, mean arterial pressure (MAP), uterine artery pulsatility index (UtA-PI), and placental growth factor (PlGF), whereas MaiRa generates an individualized molecular risk estimate from maternal plasma cfRNA profiling reflecting early biological processes at the maternal–fetal interface. (E) Primary study analyses, including receiver-operating-characteristic analysis, clinical threshold performance, patient-level risk concordance, and implementation strategies. EOPE, early-onset preeclampsia; FMF, Fetal Medicine Foundation; MAP, mean arterial pressure; PlGF, placental growth factor; UtA-PI, uterine artery pulsatility index; cfRNA, cell-free RNA.

**Table 1.**
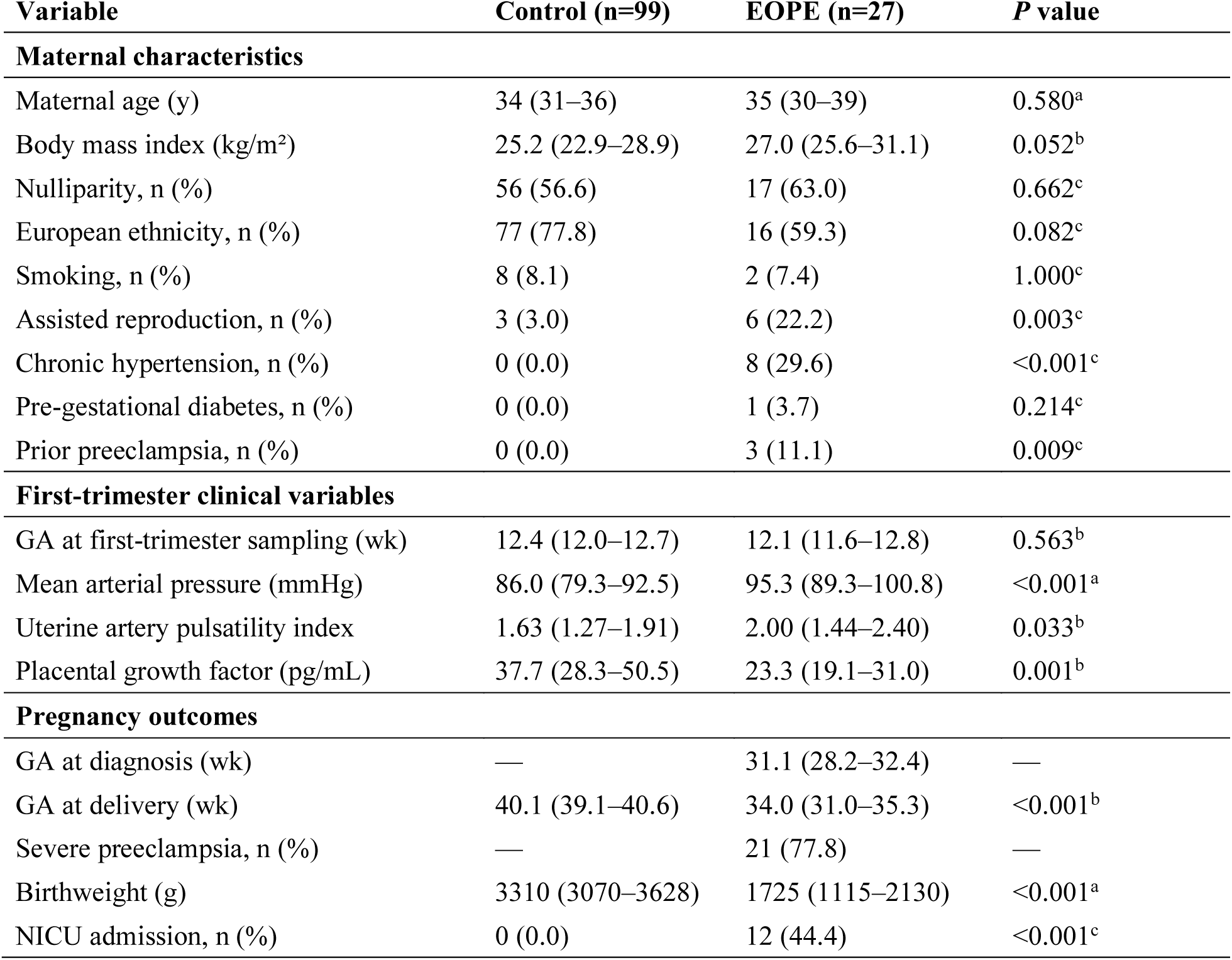
Maternal characteristics, first-trimester screening variables, and pregnancy outcomes. Values are given as medians (interquartile ranges) or absolute values (percentages). P values were calculated using ᵃ Welch’s *t* test for approximately normally distributed continuous variables, ᵇ the Mann– Whitney *U* test for non-normally distributed continuous variables, and ᶜ Fisher exact test for categorical variable. BMI, body mass index; EOPE, early-onset preeclampsia; FMF, Fetal Medicine Foundation; GA, gestational age; NICU, neonatal intensive care unit; PlGF, placental growth factor.

| Variable | Control (n=99) | EOPE (n=27) | P value |
| --- | --- | --- | --- |
| **Maternal characteristics** |  |  |  |
| Maternal age (y) | 34 (31–36) | 35 (30–39) | 0.580<sup>a</sup> |
| Body mass index (kg/m<sup>2</sup>) | 25.2 (22.9–28.9) | 27.0 (25.6–31.1) | 0.052<sup>b</sup> |
| Nulliparity, n (%) | 56 (56.6) | 17 (63.0) | 0.662<sup>c</sup> |
| European ethnicity, n (%) | 77 (77.8) | 16 (59.3) | 0.082<sup>c</sup> |
| Smoking, n (%) | 8 (8.1) | 2 (7.4) | 1.000<sup>c</sup> |
| Assisted reproduction, n (%) | 3 (3.0) | 6 (22.2) | 0.003<sup>c</sup> |
| Chronic hypertension, n (%) | 0 (0.0) | 8 (29.6) | <0.001<sup>c</sup> |
| Pre-gestational diabetes, n (%) | 0 (0.0) | 1 (3.7) | 0.214<sup>c</sup> |
| Prior preeclampsia, n (%) | 0 (0.0) | 3 (11.1) | 0.009<sup>c</sup> |
| **First-trimester clinical variables** |  |  |  |
| GA at first-trimester sampling (wk) | 12.4 (12.0–12.7) | 12.1 (11.6–12.8) | 0.563<sup>b</sup> |
| Mean arterial pressure (mmHg) | 86.0 (79.3–92.5) | 95.3 (89.3–100.8) | <0.001<sup>a</sup> |
| Uterine artery pulsatility index | 1.63 (1.27–1.91) | 2.00 (1.44–2.40) | 0.033<sup>b</sup> |
| Placental growth factor (pg/mL) | 37.7 (28.3–50.5) | 23.3 (19.1–31.0) | 0.001<sup>b</sup> |
| **Pregnancy outcomes** |  |  |  |
| GA at diagnosis (wk) | — | 31.1 (28.2–32.4) | — |
| GA at delivery (wk) | 40.1 (39.1–40.6) | 34.0 (31.0–35.3) | <0.001<sup>b</sup> |
| Severe preeclampsia, n (%) | — | 21 (77.8) | — |
| Birthweight (g) | 3310 (3070–3628) | 1725 (1115–2130) | <0.001<sup>a</sup> |
| NICU admission, n (%) | 0 (0.0) | 12 (44.4) | <0.001<sup>c</sup> |

Maternal age, body mass index, nulliparity, ethnicity, smoking status, and gestational age at first-trimester sampling did not differ significantly between groups (P>.05). Assisted reproduction (P=.003) and chronic hypertension (*P*<.001) were more frequent among pregnancies that developed EOPE. At the first-trimester risk assessment, these pregnancies had higher mean arterial pressure (P<.001), higher uterine artery pulsatility index (P=.033), and lower placental growth factor concentrations (P=.001) than controls. Among EOPE cases, the median gestational age at disease onset was 31.1 weeks; these pregnancies delivered earlier (P<.001), had lower birthweight (P<.001), and more frequently required neonatal intensive care unit admission (*P*<.001).

### Screening performance of MaiRa for first-trimester EOPE prediction

MaiRa showed higher discrimination than combined screening (AUC, 0.974 vs 0.900; ΔAUC, 0.074; 95% CI, 0.003–0.161; *P*=.040, paired bootstrap; Figure 2A). At its prespecified clinical threshold of ≥0.251, MaiRa identified 24 of 27 EOPE cases, yielding a sensitivity of 88.9% and a specificity of 90.9% (Figure 2B) whereas at the prespecified FMF risk threshold of ≤1/100, combined screening identified 15 of 27 cases, yielding a sensitivity of 55.6% and a specificity of 92.9%. (Figure 2C). Patient-level distributions showed clearer separation between EOPE and control pregnancies with MaiRa, whereas FMF showed greater overlap and left more EOPE cases classified as low risk (Figure 2B and 2C).

**Figure 2.**
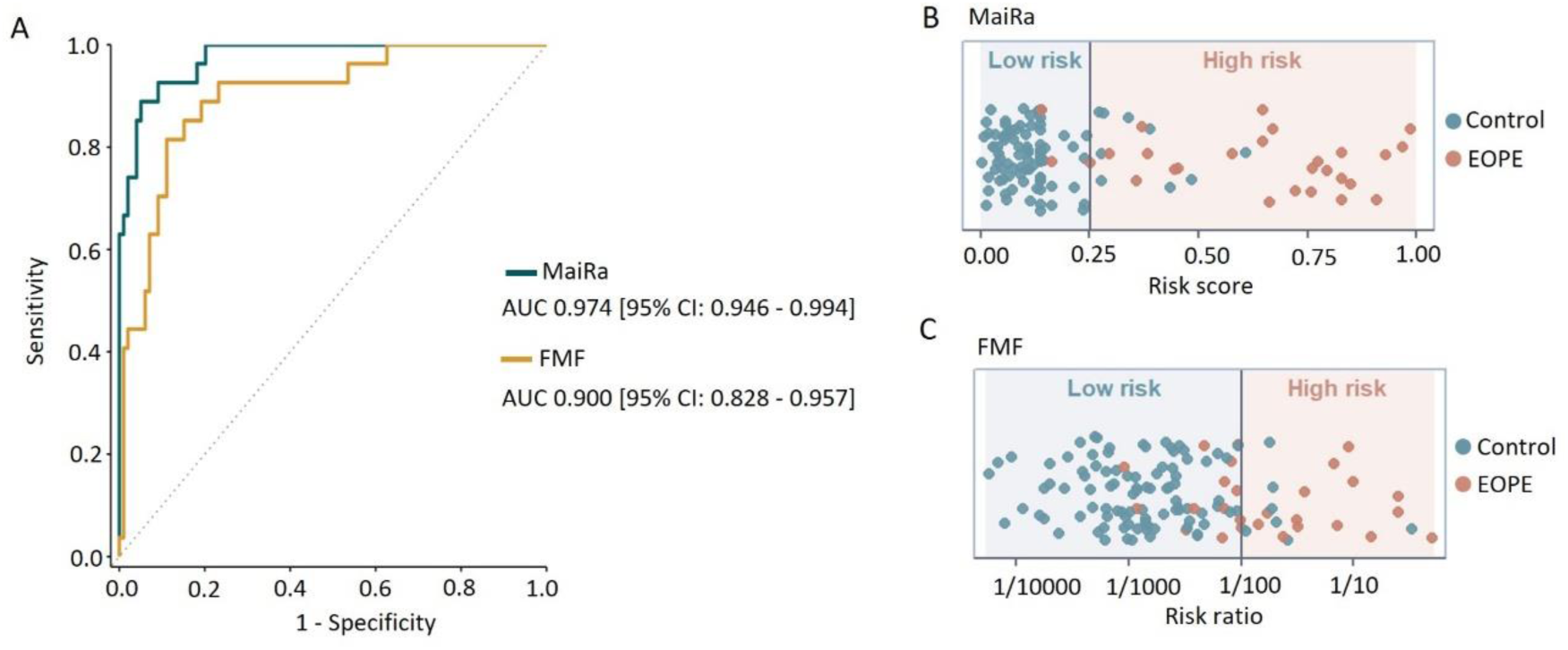
Discriminatory performance of MaiRa and FMF for first-trimester EOPE prediction. The analysis included 126 pregnancies, comprising 27 EOPE cases and 99 controls. (A) Receiver-operating-characteristic curves for MaiRa and FMF. (B–C) Patient-level distributions of MaiRa scores and FMF risk estimates at the prespecified clinical thresholds of MaiRa ≥0.251 and FMF ≤1/100. Background shading indicates the low- and high-risk regions defined by each threshold. Dot colors indicate the observed pregnancy outcome.

Across fixed false-positive rates, MaiRa consistently achieved higher EOPE detection than FMF screening (Table 2). The difference was most pronounced at lower false-positive rates; with MaiRa detecting 85.2% and 92.6% of EOPE cases at 5% and 10% false-positive rates, compared with 44.4% and 70.4% for FMF, respectively.

**Table 2.**
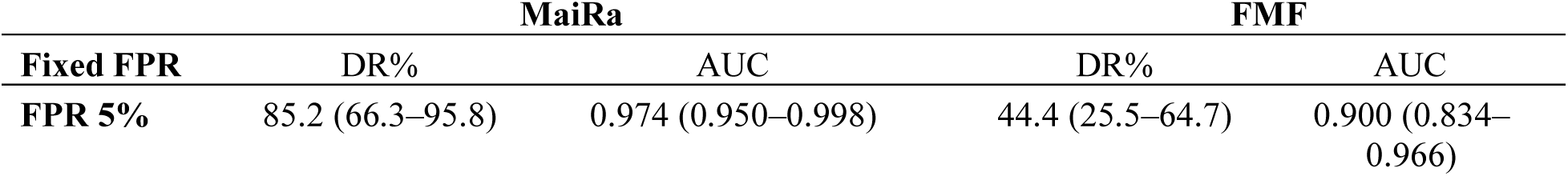
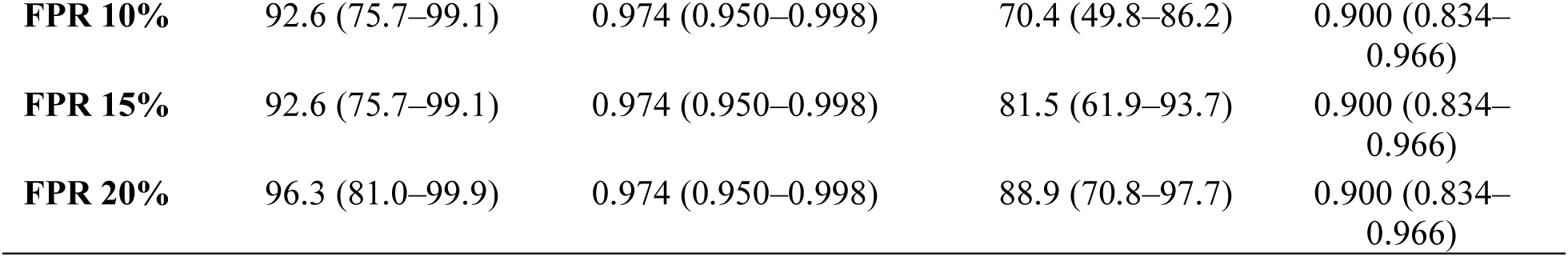
Detection performance of FMF and MaiRa for first-trimester EOPE screening at fixed false-positive rates. Detection rates and areas under the receiver-operating-characteristic curve for FMF and MaiRa at fixed false-positive rates of 5%, 10%, 15%, and 20% among 126 pregnancies, including 27 EOPE cases. Values in parentheses are 95% confidence intervals. Test-specific thresholds were selected to achieve each predefined false-positive rate. Detection rate corresponds to sensitivity. AUC, area under the receiver-operating-characteristic curve; DR, detection rate; FMF, Fetal Medicine Foundation; FPR, false-positive rate.

### Patient-level classification and clinical phenotypes

We next examined how individual pregnancies were classified by each method (Figure 3). Of the 99 controls, 85 (85.8%) were classified as low risk by both methods, 7 (7.1%) as high risk by MaiRa only, 5 (5.1%) by FMF only, and 2 (2.0%) by both methods (Figure 3, A and B). Aspirin prophylaxis had been administered in all controls classified as high risk by both methods, compared with only 1 of the 5 controls classified as high risk by FMF alone. Because prophylaxis may reduce the risk of preeclampsia in high-risk women, the absence of preeclampsia in the concordant high-risk group may partly reflect treatment-modified outcomes rather than true false-positives, although data on treatment regimen, timing, and adherence were unavailable.

**Figure 3.**
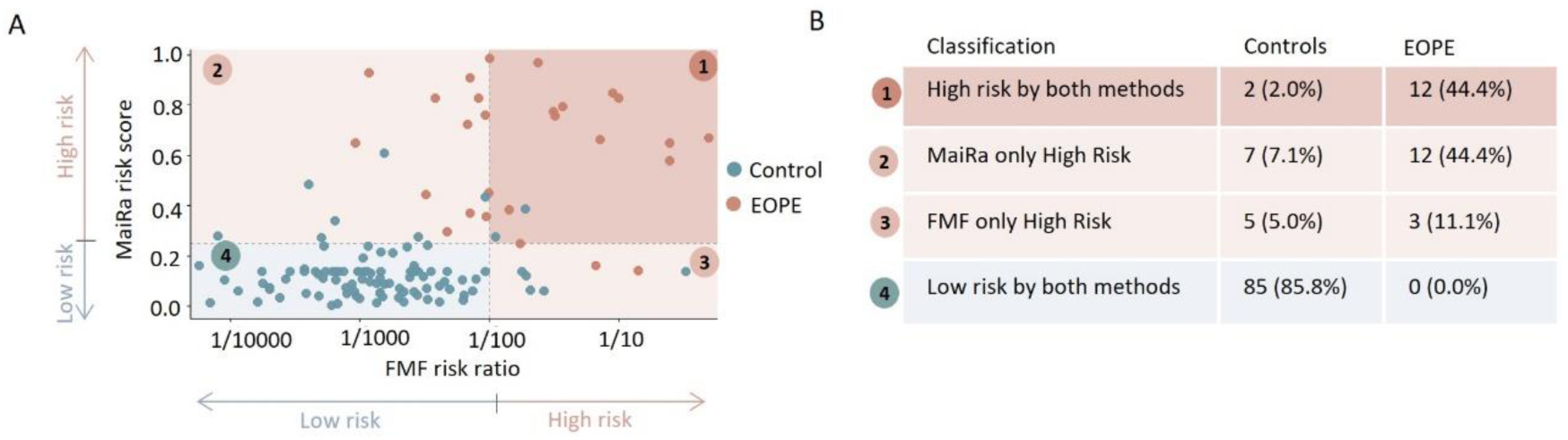
Patient-level concordance of EOPE risk classification by MaiRa and FMF. (A) Cross-classification of pregnancies according to the prespecified clinical thresholds for MaiRa (≥0.251) and FMF (≤1/100). Background shading indicates the four resulting risk regions: high risk by both tests, high risk by MaiRa only, high risk by FMF only, and low risk by both tests. Dot colors indicate the true clinical outcome. (B) Summary of patient-level classification according to MaiRa and FMF, stratified by clinical outcome. Values are presented as number and percentage of controls or EOPE cases classified as high risk by both methods, high risk by MaiRa only, high risk by FMF only, or low risk by both methods.

Among the 27 EOPE pregnancies, 12 (44.4%) were classified as high risk by both methods, 12 (44.4%) by MaiRa only, and 3 (11.1%) by FMF screening only; none was low risk by both (Figure 3, A and B). Classification discordance was therefore asymmetric, with substantially more EOPE pregnancies identified exclusively by MaiRa than exclusively by FMF screening. Consistent with the patient-level and phenotypic analyses, agreement between the two binary classifications was fair at the prespecified FMF high-risk threshold of ≤1/100 (Cohen’s κ=0.38; 95% CI, 0.19–0.56; crude agreement, 78.6%) and moderate at the broader ≤1/300 cutoff (κ=0.45; 95% CI, 0.27–0.61; crude agreement, 76.2%). Rather than reflecting measurement discordance, this partial agreement indicates that MaiRa and FMF capture distinct, non-redundant high-risk profiles, reinforcing the added value of molecular screening for identifying EOPE pregnancies that conventional markers leave below the risk threshold.

To characterize the EOPE pregnancies identified by MaiRa but missed by FMF, we examined maternal risk factors, conventional first-trimester screening markers, and subsequent disease severity (Figure 4A). Pregnancies flagged as high risk by both methods generally showed a greater burden of maternal risk factors and more pronounced abnormalities in mean arterial pressure and uterine artery pulsatility index. In contrast, those identified exclusively by MaiRa had fewer maternal risk factors and less abnormal conventional screening profiles, with similar placental growth factor values. These findings indicate that EOPE pregnancies identified exclusively by MaiRa exhibited less abnormal conventional screening profiles despite developing clinically severe disease (Figure 4B). Despite their less abnormal conventional risk profiles, 8 of the 12 pregnancies identified exclusively by MaiRa subsequently developed severe EOPE, indicating that the additional cases detected by MaiRa were not restricted to clinically milder disease.

**Figure 4.**
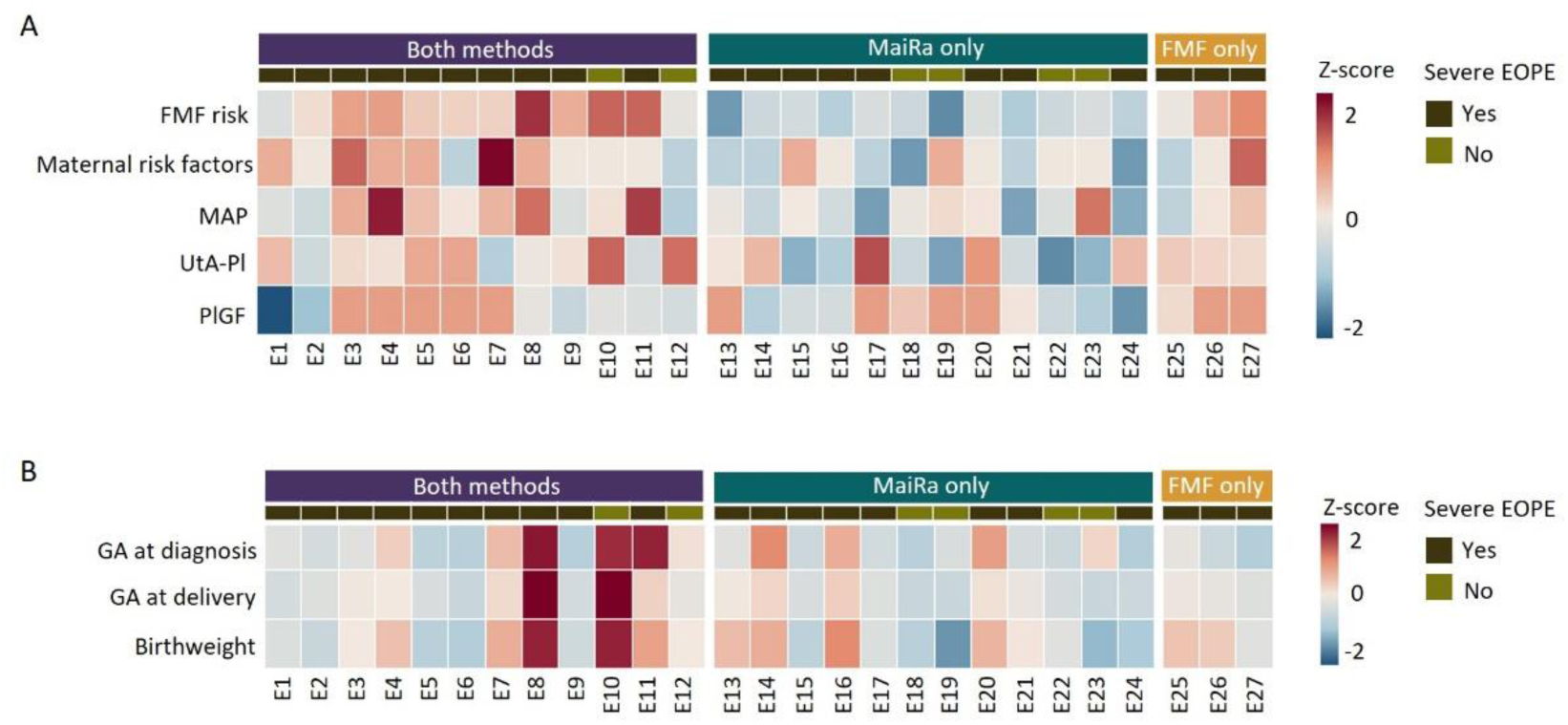
Clinical phenotype of EOPE pregnancies identified by MaiRa and FMF. (A) Heatmap of the 27 EOPE cases, grouped according to whether they were classified as high risk by both tests, by MaiRa only, or by FMF only. Rows show FMF risk, maternal risk burden, mean arterial pressure, uterine artery pulsatility index, and placental growth factor. Maternal risk burden was calculated as the unweighted sum of 8 binary maternal risk factors: maternal age ≥35 years, obesity, previous preeclampsia, nulliparity, chronic hypertension, pregestational diabetes, autoimmune disease, and Afro-American ethnicity. Continuous variables were standardized as z scores across EOPE cases. PlGF values were directionally reversed before standardization so that positive z scores and red shading consistently indicate a more adverse conventional risk profile. Top annotation bars indicate classification group and the presence or absence of severe EOPE. (B) Heatmap of gestational age at diagnosis, gestational age at delivery, and birthweight in the 27 EOPE cases, grouped as high risk by both tests, by MaiRa only, or by FMF only. Continuous variables were standardized as z scores across EOPE cases. Lower values indicate earlier diagnosis, earlier delivery, and lower birthweight, respectively. Top annotation bars indicate classification group and the presence or absence of severe EOPE.

### Potential implementation pathways for MaiRa-based molecular screening

Having demonstrated the diagnostic performance of MaiRa as a standalone molecular screening test for EOPE, we explored two potential implementation models using the prespecified clinical threshold (≥0.251): universal molecular screening and a contingent strategy in which conventional FMF screening was used to triage pregnancies for subsequent MaiRa testing (Table 3).

**Table 3.** Performance of potential implementation strategies for first-trimester molecular EOPE screening. Performance of three potential implementation models for first-trimester EOPE screening among 126 pregnancies (99 controls and 27 EOPE cases). In the contingent strategy, pregnancies with an FMF risk of ≤1/300 underwent subsequent MaiRa testing using the prespecified clinical threshold of ≥0.251; whereas pregnancies above the FMF triage threshold were classified as low risk without molecular testing. MaiRa testing indicates the number and proportion of pregnancies requiring molecular analysis.

|  | **FMF standalone** | **Universal MaiRa** | **Contingent MaiRa** |
| --- | --- | --- | --- |
| Screening approach | FMF $\leq 1/100$ | MaiRa $\geq 0.251$ | FMF $\leq 1/300$ followed by MaiRa $\geq 0.251$ |
| Sensitivity, % | 55.6 | 88.9 | 77.8 |
| Specificity, % | 92.9 | 90.9 | 97.0 |
| MaiRa testing, n/N (%) | 0/126 (0) | 126/126 (100) | 45/126 (35.7) |

FMF standalone identified 55.6% of EOPE cases with 92.9% specificity. Universal MaiRa screening provided the highest detection observed in this cohort (88.9% of cases; 90.9% specificity), with molecular testing in all pregnancies. The contingent strategy (FMF triage ≤1/300 followed by MaiRa) achieved 77.8% sensitivity and 97.0% specificity while limiting molecular testing to 35.7% of pregnancies.

These exploratory analyses illustrate that MaiRa may be deployed either as a universal screen, maximizing detection, or, where universal cfRNA testing is not yet feasible, within a contingent pathway that substantially reduces testing volume while preserving high performance.

## Comment

### Principal findings

MaiRa showed greater first-trimester discrimination for EOPE than the FMF algorithm when both approaches were evaluated in the same pregnant individuals. Patient-level analyses indicated that the two methods did not simply rank the same pregnancies differently but identified partially distinct risk profiles. In particular, MaiRa detected EOPE cases that were classified as low risk by FMF and that generally showed fewer maternal risk factors and less pronounced abnormalities in conventional first-trimester markers, despite including pregnancies that subsequently developed severe disease. This discordant profile is consistent with the recognized heterogeneity of preeclampsia and suggests that clinically relevant EOPE biology may not be fully captured by maternal characteristics, biophysical measurements, and circulating protein markers alone. Exploratory clinical-use analyses further showed that MaiRa could be implemented either as a standalone screening test or within a contingent pathway designed to reduce molecular-testing burden.

### Results in the context of what is known

First-trimester combined screening with the FMF algorithm, followed by aspirin prophylaxis in women classified as high risk, is an established strategy for reducing preterm preeclampsia, particularly in Europe.^3,9^ However, recent population-based data suggest that its impact may be more limited in routine practice.^37^ In a population-screening setting, even modest reductions in specificity can translate into substantial numbers of unaffected women being classified as high risk. Improving EOPE detection while limiting false-positive classifications therefore remains a central challenge, because greater sensitivity with conventional screening generally requires accepting a higher false-positive rate.^11,13^

Maternal plasma cfRNA profiling offers a broader molecular representation of pregnancy biology than conventional first-trimester screening by capturing dynamic transcriptional activity associated with maternal and placental adaptation. Recent multimodal cell-free DNA studies have likewise demonstrated the potential of circulating nucleic acid profiling for first-trimester prediction of preeclampsia, supporting a broader shift toward molecular screening approaches in obstetric medicine.^38^ Whereas those approaches integrate fragmentomic, epigenetic, and clinical features, MaiRa is based on maternal plasma cfRNA, providing a direct molecular readout of dynamic transcriptional activity associated with early disease biology. In the present head-to-head comparison, MaiRa showed greater discrimination than FMF, particularly when high sensitivity was prioritized. Our patient-level analyses suggest that these differences are unlikely to reflect simple changes in ranking performance. Rather, they indicate that the two approaches interrogate partially distinct aspects of EOPE biology.

Pregnancies identified exclusively by MaiRa generally showed fewer maternal risk factors and less pronounced abnormalities in MAP and UtA-PI than cases identified by both methods, despite broadly similar PlGF profiles and a high frequency of severe disease. This pattern is consistent with the biological heterogeneity of preeclampsia.^28–30^ Rather than representing milder disease, many progressed to severe EOPE despite exhibiting less abnormal conventional screening profiles. This observation supports the concept that different biological pathways may converge toward the same clinical syndrome. By capturing dynamic transcriptional signals from maternal and placental tissues,^31–35^ the circulating transcriptome may reflect multiple disease pathways simultaneously and identify biological changes that are not yet sufficiently represented by conventional first-trimester screening variables.

These differences may also relate to the endpoint used for model development. The FMF algorithm estimates the risk of preeclampsia leading to delivery before 37 weeks, whereas MaiRa predicts disease onset before 34 weeks. Because delivery timing is influenced by fetal condition, surveillance, local practice, and obstetric intervention, a delivery-based endpoint may incorporate management-related variability.^39,40^ By targeting disease onset, MaiRa may reduce this source of noise and align prediction more closely with the underlying biological process.

### Clinical and research implications

The observation that MaiRa identified EOPE pregnancies that remained below conventional screening thresholds has implications beyond comparative test performance. Rather than relying primarily on maternal characteristics and downstream biophysical or biochemical manifestations of placental dysfunction, maternal plasma cfRNA profiling provides a direct molecular readout of decidual function within a broader landscape of placental, vascular, and maternal immune processes underlying pregnancy adaptation.. If confirmed in larger prospective studies, molecular screening may therefore represent a complementary paradigm for first-trimester EOPE risk assessment, capable of identifying clinically relevant disease that is not fully captured by current screening approaches.

From a clinical perspective, the exploratory implementation analyses illustrate that MaiRa could be deployed through different screening pathways according to local priorities and available resources. Universal molecular screening provided the highest EOPE detection while maintaining high specificity. Where universal cfRNA testing is not yet feasible, a contingent strategy using FMF for initial triage substantially reduced the number of pregnancies requiring molecular analysis, although its sensitivity remained constrained by the initial selection of pregnancies for testing. These approaches should therefore be regarded as alternative implementation models whose relative value will depend on clinical objectives, infrastructure, and cost-effectiveness.

MaiRa also offers practical advantages for routine implementation. Conventional combined screening integrates maternal characteristics with biochemical assays and operator-dependent biophysical measurements—mean arterial pressure and uterine artery pulsatility index—that are subject to inter-observer variability, so performance may differ across sonographers and centers. In contrast, MaiRa derives from a cfRNA signature measured in a single maternal blood sample using a standardized, automated laboratory workflow, minimizing operator dependence and favoring reproducibility across centers. This may broaden access to standardized first-trimester risk assessment, particularly where high-quality uterine artery Doppler or complete combined screening is not consistently available, and supports a universal strategy that captures EOPE cases misclassified as low risk by conventional screening.

Beyond first-trimester EOPE screening, the longitudinal nature of circulating cfRNA suggests that molecular risk assessment could evolve throughout pregnancy. Future prospective studies should determine whether serial cfRNA profiling improves prediction across gestational windows, enables dynamic risk reassessment, or supports individualized surveillance and preventive strategies.

### Strengths and limitations

This study has several strengths. The nested case-control design within a prospective, multicentre cohort allowed a direct, patient-level comparison of MaiRa and FMF combined screening performed in the same first-trimester pregnancies, avoiding the between-population variability that limits indirect test comparisons. Discrimination was compared using a paired bootstrap procedure that accounted for the correlated nature of the two measurements, and the analysis extended beyond summary performance metrics to characterize discordant risk phenotypes at the individual level and to evaluate alternative implementation models.

Several limitations should be acknowledged. The sample was modest and enriched for cases through a case-control design, which inflates disease prevalence relative to a screening population and precludes direct estimation of positive predictive value or screen-positive rates under real-world conditions. Cohen’s κ is also sensitive to disease prevalence; because the cohort was enriched for cases, the absolute κ values should be interpreted as an internal comparison between methods rather than as agreement estimates directly transferable to an unselected screening population. MaiRa was evaluated in the same cohort in which it was developed and internally validated, so its apparent performance may be optimistic and requires external confirmation. The implementation scenarios were exploratory and based on small numbers, the FMF triage cutoffs were derived post hoc within this dataset, and information on aspirin use, treatment timing, and adherence was incomplete, which may have modified outcomes among high-risk pregnancies. Finally, the analysis was restricted to singleton pregnancies with complete first-trimester data and did not include a health-economic evaluation of molecular screening. Accordingly, the comparative performance and the proposed implementation strategies require prospective confirmation in an independent, unselected obstetric population before clinical adoption.

### Conclusions

MaiRa improved first-trimester discrimination of EOPE compared with the FMF algorithm and identified EOPE pregnancies that were not captured by conventional screening, including pregnancies that subsequently developed severe disease. These findings suggest that maternal plasma cfRNA profiling may capture clinically relevant biological processes not fully represented by conventional first-trimester screening and support further prospective evaluation as a molecular approach to EOPE screening.

## Data Availability

Data supporting the findings of this study are available from the corresponding authors upon reasonable request, subject to applicable ethical and data-protection restrictions.

